# Ability of Fabric Facemasks Materials to Filter Ultrafine Particles at Coughing Velocity

**DOI:** 10.1101/2020.04.14.20065375

**Authors:** Eugenia O’Kelly, Sophia Pirog, James Ward, P. John Clarkson

**Affiliations:** Department of Engineering, University of Cambridge – currently in San Francisco, USA; Department of Medical Social Sciences, Northwestern University – Chicago, USA; Department of Engineering, University of Cambridge – Cambridge, UK

**Keywords:** SARS-CoV-2, Coronavirus, Infection Control, Respiratory Infections, Facemask, Public Health, Infectious Disease, PPE

## Abstract

**Objectives:** We examined the ability of fabrics which might be used to create homemade face masks to filter out ultrafine (0.1μm and smaller in diameter) particles at the velocity of adult human coughing.

**Method:** Twenty commonly available fabrics and materials were evaluated for their ability to reduce air concentrations of ultrafine particles at a face velocity of 16.5 m/s. Further assessment was made on the filtration ability of select fabrics while damp and of fabric combinations which might be used to construct homemade masks.

**Results:** Single fabric layers blocked a range of ultrafine particles. When fabrics were layered, significantly more ultrafine particles were filtered. Nonwoven fusible interfacing significantly increased filtration.

**Conclusions:** The current coronavirus pandemic has left many communities without access N95 facemasks. Our findings suggest that face masks made from layered common fabric can help filter ultrafine particles and provide some protection for the wearer when commercial facemasks are unavailable.

**STRENGHTS AND LIMITATIONS OF THIS STUDY:**

- Tested a large number of potential facemask materials, including materials currently in common use such as Lycra which have not been previously tested
- Evaluated filtration efficiency at coughing velocities, more closely mimicking use-case of masks worn for community protection than previous studies
- Assess the data from prior published work and current study, creating a picture of Filtration Efficiency and the impact of velocity
- Did not discriminate between pathogenic and non-pathogenic particles
- Breathing resistance was estimated based on qualitative feedback

## INTRODUCTION

The current SARS-CoV-2 outbreak has left many communities without sufficient quantities of face masks for the protection of medical staff and first responders, let alone sufficient quantities of masks for the general population’s use. Despite this severe shortage, many areas have begun requiring the use of facemasks for individuals who leave their property. Both the United Kingdom and United States have begun mandating the of fabric face masks for individuals while many scholars continue strongly encourage the precautionary use of face coverings[1].

Homemade face masks have now become a necessity for many to both meet the demands that cannot be met by supply chains and/or to provide more affordable options. Although widespread online resources are available to help home sewers and makers create masks, scientific guidance on the most suitable materials is currently limited.

Though not as effective as surgical masks or respirators, homemade face masks have been shown to provide benefit in filtering viral and bacterial particles[2-4]. The primary purpose of face masks worn by the general public is to limit the spread of viral particles from respiratory activity, rather than blocking the inhalation of any contagious particles[5]. For the protection of the face- mask wearer, the Center for Disease Control specifically recommends fabric face masks for the purpose of limiting viral spread through respiratory droplet[5,6]. Face masks worn for the protection of others must efficiently filter particles emitted while coughing, when large amounts of potentially infectious respiratory droplets are produced.

Prior studies evaluating the efficacy of fabric face masks have tested their filtration ability under velocities representative of normal to active breathing[2-4]. Significantly more potentially infectious particles are generated and spread by coughing, which occurs at velocities up to 100 times greater than those tested in previous experiments[7,8]. This study evaluates the effectiveness of fabrics to filter ultrafine particles at velocities representative of adult coughing. Although no previous studies have evaluated the ability of face masks to filter particles at high velocities, evidence suggested high velocities may significantly decrease the efficacy of face mask materials[9,10].

Furthermore, past studies have tested a limited set of similar materials, namely t-shirts, sweatshirts, scarves, and tea towels. Communicating with the international community of home sewers and small businesses seeking to design face masks, we determined a need for the assessment of a much wider range of fabric types, including stretch fabrics, felts, wool, and nylon. Some fabrics, such as stretch Lyrics and nylon, are in frequent use in commercial and homemade face masks but have not been evaluated for filtration efficiency. Conversations with material scientists and sewers highlighted the need to consider the possible benefits of nonwoven interfacing, a material not previously tested for filtration.

Finally, our study assesses the impact of moisture, an effect of respiration, on filtration efficiency. A selection of fabrics was tested when damp to simulate dampness from sweat or heavy respiration. Furthermore, as fabric face masks are often washed and re-worn, we tested all materials after subjecting them to one cycle in a home laundry machine. The literature evaluating the impact of washing and drying of fabric face masks is limited. One study on one fabric face mask showed a decrease in filtration efficiency with washing[11]. All fabric materials were tested after one wash and dry in a home machine.

Both individual materials and material combinations were tested with the goal of increasing particle filtration of homemade masks.

## METHODS

This study was conducted in response to the rapidly growing SARS-CoV-2 outbreak. As such, priority was given to developing a test apparatus which could be constructed and provide usable results in a short amount of time.

### Patient and Public Involvement

The research team communicated closely with home sewers, small businesses branching out to include fabric face mask manufactures, and physicians interested in protecting at-risk patients when masks were not available. Our conversations highlighted a need for filtration information on a wider variety of materials than those assessed in previous studies. We studied a range of materials that were previously unexamined in the literature, but of high interest to the aforementioned communities. These included: felt, Lycra, felts, washable vacuum bags, and quilt batting/wadding. Materials for investigation were selected based on those that home sewers reported as being readily available. Responding to home sewers’ understanding of fabric categories and the success of cotton in prior research[2-4], we also tested various weaves of cotton commonly available, including quilting cotton, shirting cotton, and cotton jersey knit.

The physician and home sewing communities raised concerns regarding the risks of infection by reusing masks. In response to this, preference was given to materials which could be cleaned in a home washing machine and/or dryer at its hottest setting. All materials were washed and dried before testing. This caused significant shrinkage of wool felt. In response to further concerns about efficacy when damp, top-performing materials were subjected to five additional tests when damp.

### Testing Apparatus

Tests were conducted as described by Hutten[12]. An airtight apparatus allowed simultaneous testing of unfiltered and filtered air. A 2.5 cm diameter tube provided access to two ultrafine particle counters (P-Trak model 8525) which measured concentrations of particles 0.1 μm and smaller. The tube held a 2.5 cm diameter sample of the filter material. Material was allowed to relax on a flat surface and the testing mount placed on top, with excess material secured by an adjustable clip. See Figure 1 for an illustration of the testing apparatus.

**Figure 1:**
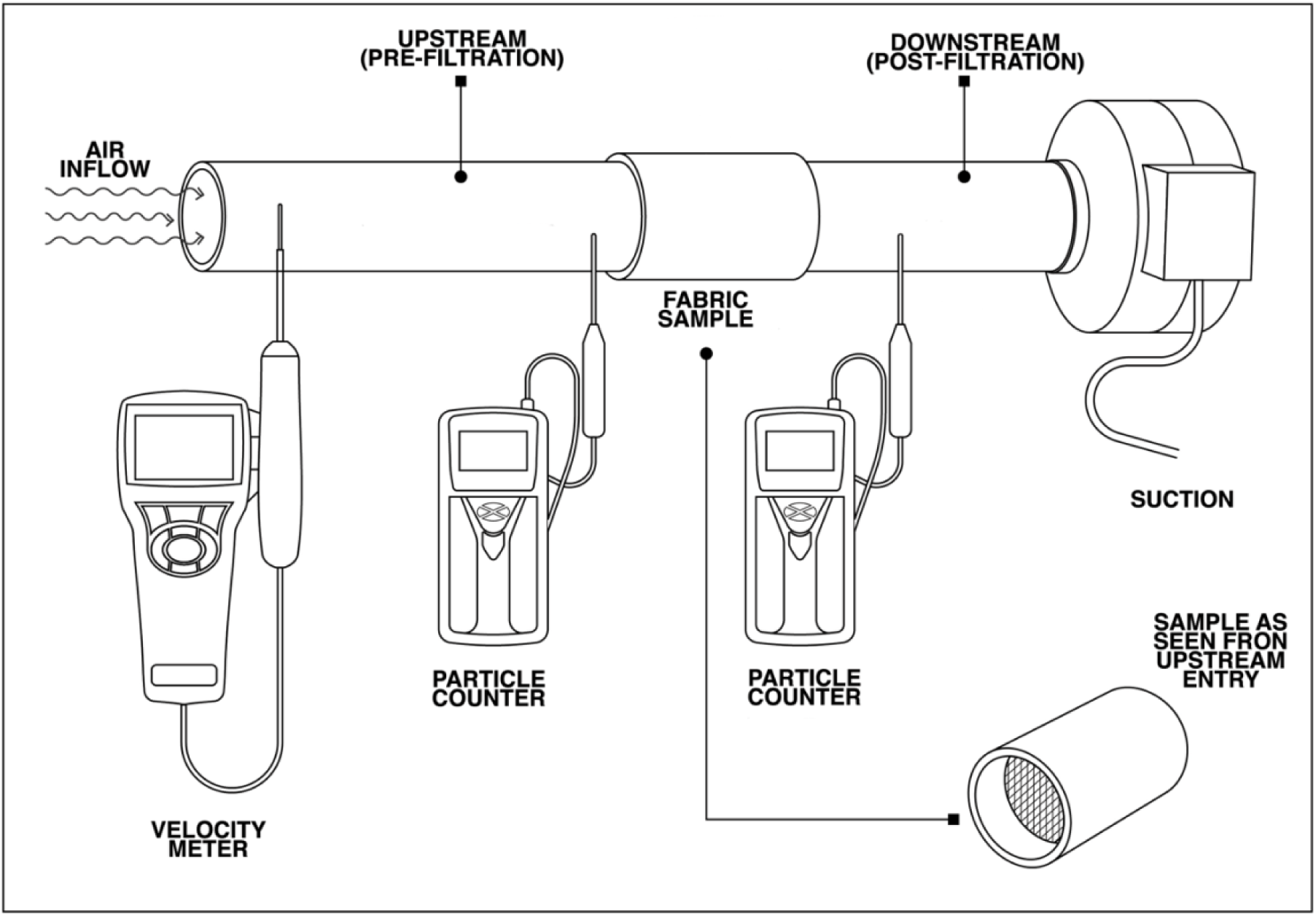
Diagram of experimental apparatus using two P-Trak Ultrafine Particle 8525 counters for simultaneous measurement and a TSI 9565 VelociCalc to measure face velocity.

After mounting a new specimen, a minimum of three minutes loading time at high velocity was given. At least thirty seconds between sequential tests on a previously loaded material was given.

Probes for the velocity meter and particle counters were inserted halfway into the tube. Flexible sealant was used around the entry points of the probes to prevent air leakage.

Airflow was controlled through suction, which pulled air through the filter medium at a rate of approximately 16.5 m/s. This number was chosen as a median between the average face velocity (11.2 m/s) and greatest face velocity (22 m/s) recorded in a study on saliva droplet transport by adult coughing[7]. Face velocity represents the speed of the particles when leaving the mouth. The chosen velocity was also in line with the 15.3 m/s average initial coughing velocity of an adult male measured in a 2012 study[13].

Prior to conducting high velocity tests, a calibration test was performed to validate the testing apparatus at low velocities. Five control tests at low velocity (suction placed 20 cm distance from downstream air intake) showed the N95 performance averaging 89% with a high of 93%. A high-quality PM 2.5 filter showed an average FE of 89% and a high of 90%. Velocity for this calibration test was not recorded.

### Calculating Filtration Efficiency

Filtration efficiency represents the percent of particles a filter medium can block. Hutten’s formula was used to assess filtration efficiency (FE).

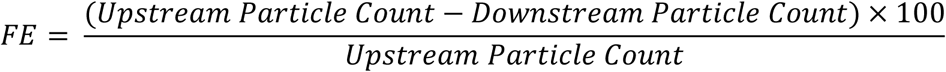

For each material or material combination, ten sets of readings were collected. Readings were collected using two P-Trak Ultrafine Particle Counters, Model 8525. Each reading was collected as a 10-second average of ultrafine air particle concentrations. The average filtration efficiency for each material was calculated. Due to the number of readings collected, the 95% confidence intervals for error bars was calculated using the appropriate *t* distribution critical value.

### Breathing Resistance

To estimate the breathing resistance of each material, and thus their suitability for use in a face mask, two members of the team held sections of each fabric tightly over their mouth and inhaled through their mouth. Each fabric was scored on a 0-3 scale where 3 represented a great difficulty in drawing breath, 2 represented that there was noticeable resistance, but breath could be drawn, 1 represented minor limitation but relative ease of breathing, and 0 represented no noticeable hindrance. Combining and layering fabric was not found to significantly increase the breathing difficulty. All face mask fabric combinations scored 1 or 2.

### Damp Testing

Dampness was achieved by applying 7 milliliters of filtered water, the approximate amount of water exhaled by an adult during an hour of respiration[14], to the 5 cm square section of the material.

### Note on Study Design and Limitations

It should be noted that, due to the limitations imposed by this outbreak, this study was done with available materials. Data from this study should be treated as preliminary and used to inform decisions about filtration media only in relation to existing studies which assess viral filtration through the collection of viral cultures.

Ten readings were taken for each material, although one reading for the disposable HEPA vacuum bag had to be later discarded due to a data transfer error. At least two different sections of each type of fabric were tested to ensure accurate representation of the material. Zero readings were taken on the particle testers regularly to ensure proper functioning.

## RESULTS

### Materials

All materials blocked some ultrafine particles (see Figure 2). A 3M N95 mask and hospital-grade surgical mask were tested for the sake of comparison. Two types of vacuum bag, a disposable HEPA vacuum bag and a washable HEPA vacuum bag, were evaluated due to the number of people attempting to utilize these materials as face mask filters. Eighteen fabrics were tested as a single layer. Lastly, fabrics were layered to represent potential mask designs. For this test, fusible interfacing was heat bonded to another layer.

**Figure 2:**
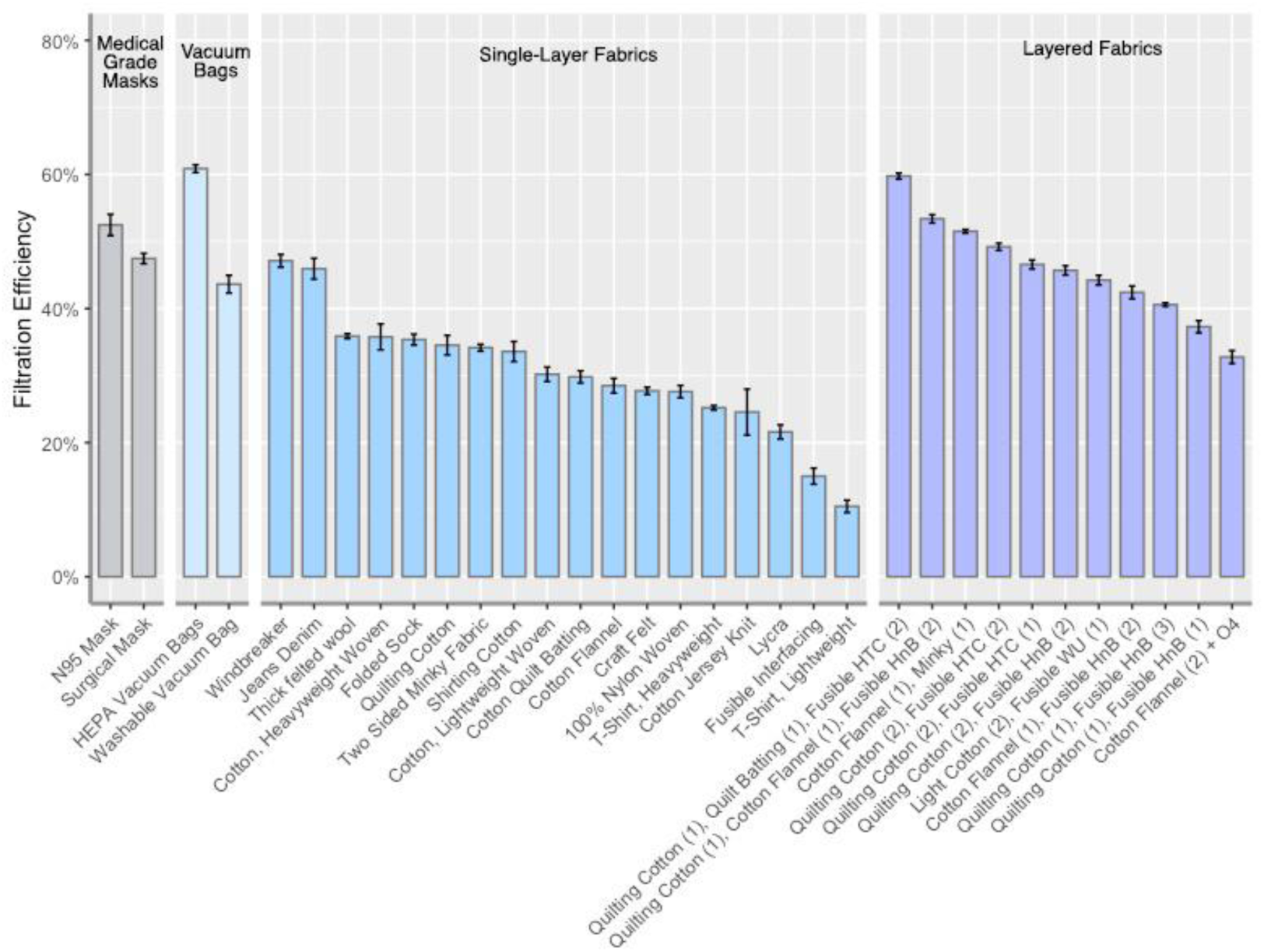
The filtration efficiency of tested fabrics and fabric combinations with error bars showing 95% confidence.

HEPA vacuum bags blocked the most ultrafine particles, with the N95 mask from 3M blocking the second greatest percentage of particles.

Repurposing HEPA filters holds great promise for emergency facemasks; however, great care should be taken that the component materials within the filter do not pose dangers to those making or wearing the face mask. While the single-use HEPA vacuum bag tested showed the greatest ability to filter ultrafine particles, the layers fell apart when the material was cut, exposing inner layers of the fabric. Vacuum bags may have component materials which are effective at filtering particles but which are unsafe to inhale or come into close contact with the face. The reusable, washable HEPA bags had a construction more suitable to creating emergency facemasks as the material held together well and did not expose inner fibers, but the safety of the materials used are also unknown.

The filtration efficiencies of select materials were tested when damp (see Figure 3). Only minor differences in filtration efficiency were noted for quilting cotton, cotton flannel, and craft felt. Denim showed a significant decrease in efficiency while the HEPA single-use vacuum bags showed an increase in efficiency when damp.

**Figure 3:**
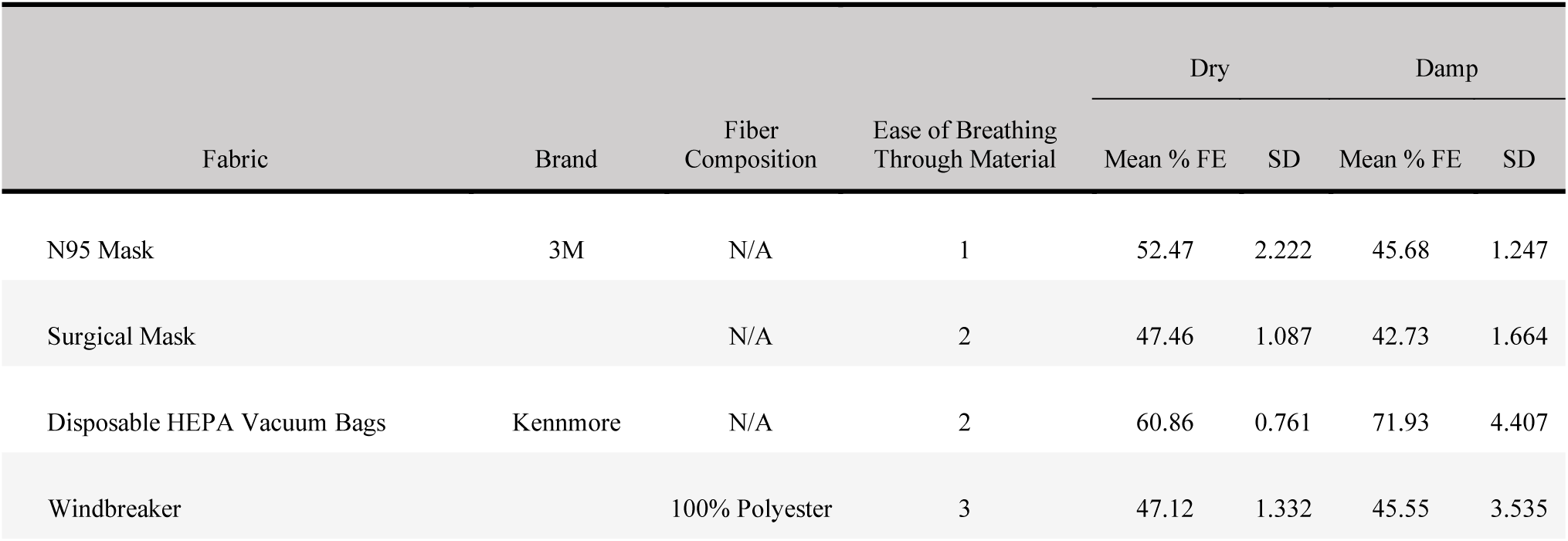
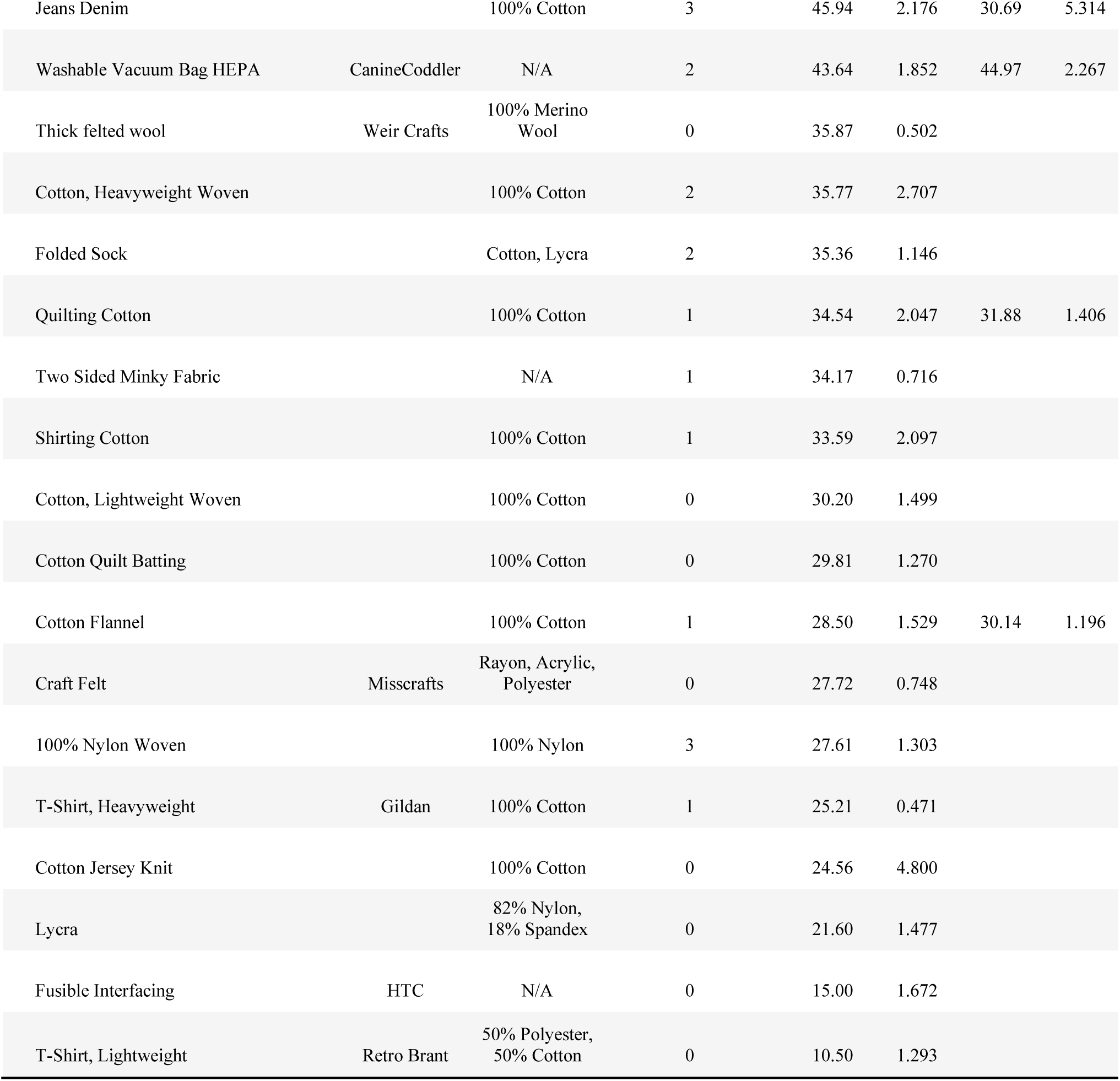
Chart of materials composition, breathing resistance, mean FE, standard deviation of FE, and, where available, FE when damp.

Figure 3 also provides breathing resistance, fabric composition, FE, and standard deviation. The most suitable fabrics for face masks are those with a high FE but low breathing resistance. Denim jeans and windbreaker fabric blocked a high proportion of ultrafine particles but were extremely difficult to breathe through (see Figure 3). The windbreaker fabric may be suited to a loose-fitting face mask which protects the wearer from liquid droplets or splashes but is unsuitable for filtration.

Suitable materials which showed high filtration efficiency and low breathing resistance included felted wool, quilting cotton, and cotton flannel. A single sock held flat compared well with the quilting cotton and, when pressed tight against the nose and mouth, may provide emergency protection.

### Nonwoven Fusible Interfacing

Nonwoven fusible interfacing, the kind used for stiffening collars and other areas in garments, was able to significantly improve the ability of the fabrics to filter ultrafine particles without increasing breathing resistance. Of particular note, we found that brands exhibited significant differences in filtering performance. HTC brand lightweight interfacing was more effective than Heat-n-Bond brand lightweight interfacing. Applying two layers of the Heat-n-Bond achieved similar improvements to filtration efficiency as the HTC brand. Wonder Under, a double sided, heavyweight fusible interfacing for constructing bags and craft projects, showed similar filtration ability to the HTC brand but may be too stiff to be suitable for face mask construction.

### Material Combinations

When layered to create potential face mask configurations, common fabrics were able to achieve much higher levels of ultrafine particle filtration (see Figure 2). Some material combinations were able to filter out higher percentages of ultrafine particles than the surgical or N95 masks tested, although this should not be taken to mean they provide higher levels of protection from viruses. All fabric combinations scored between a 2 and 3 on the breathing resistance test, indicating they were more difficult to breathe through than an N95 mask.

## DISCUSSION

Our data suggests that, in times of severe supply shortage, common fabrics can be layered to create face masks which protect wearers and others from a significant percentage of ultrafine particles. It should not be inferred that these layered fabrics can protect wearers from more viral particles than N95 masks or surgical masks as our study did not discriminate between viral particles and other ultrafine particles. Many viruses are carried on droplets or other particles significantly larger than those in tested here. Furthermore, these results do not incorporate the challenges of achieving fit, a critical factor of facemask design. The benefit of using materials which offer high filtration efficiency are likely to be significantly reduced or negated if the mask is worn with a poor fit.

Many viruses are carried on droplets or other particles significantly larger than those in tested here. Previous studies have shown that large particles are more readily filtered[3,4] than smaller particles, indicating that a study of ultrafine particles will lead to a low ‘baseline’, upon which filtration efficiency of larger particles will increase. Moreover, ultrafine particles tend to pose high risks during other emergency situations when fabric face masks are needed, such as forest fire outbreaks and times of high, concentrated pollution.

### The Effect of Velocity on Filtration Efficiency

The flow rate of air used in this study represents the velocity of air expelled during human coughing[7] and is the first such study to evaluate fabric filtration under high velocities. A velocity of 16.5 m/s or 1650 cm/s was chosen to represent the face velocity of an adult coughing[7]. N95 filtration efficiency of NaCL was seen to decrease with velocity in prior filtration studies, from 99% in Rengasamy et all’s evaluation at 0.165 m/s to 85% in Konda et all’s evaluation at 0.26 m/s. As the velocity was up to 100 times greater than Rengasmay et all’s and 63 times great than Konda eta all’s, filtration efficiency was expected to be significantly lower if velocity impacts filtration efficiency. Our results support the idea that velocity has a significant impact on filtration efficiency.

Popular mask filtration which specify a face velocity include FDA-PFE, and ASTM-PFE and utilize velocities ranging from 0.5 to 25 cm/sec. Several testing methods do not specify a face velocity but instead provide flow rate for particle generation. While face velocity cannot be derived from flow rate, the flow rates utilized in these methods of 85 L/min in the NIOSHE NaCl test and 28.3 L/min in the ASTM-BFE test are lower than Konda et all’s upper flow rate of 90 L/min, which corresponded to a face velocity of 0.26 m/s.

No prior studies have evaluated the ability of N95 face masks to filter particles at such high face velocities which can be used as a direct comparison. Rangasamy’s 2015 study on synthetic blood penetration of N95 masks found that the number of respirator samples which failed the blood penetration test increased with increasing test velocities[10]. This, along with our findings, indicates a strong need to further evaluate mask filtration at high velocities. While a leak around the downstream testing port could lead to a lowered particle count, the possibility of low performance at high velocities should be eliminated through further study.

### Comparing Fabric Filtration Efficiency

Although the results from higher velocity tests are significantly lower than previous tests, the shape of the data remains highly consistent with prior studies. The average velocity used in prior studies is 0.20 m/s, which is 82.5% of the velocity used in the prior study. When the values for high velocity filtration are increased by 82.5%, the data compares closely with data from previous research. Figure 4 compares data from studies which examine fabric filtration. Where applicable, the data chosen represented similar particle size filtration and the highest velocities offered. It should be noted that each test utilizes different methods of testing filtration efficiency and different brands of materials. Konda et all applies a maximum face velocity of 0.26 m/s utilizing NaCL aerosol (approximately 0.74 μm). Rengasamy et all similarly uses aerosolized NaCL at the lowest face velocity of 0.165 m/s. Davies et all assesses the filtration of Bacteriophage MS2 (0.023 μm) at a face velocity of 0.2 m/s. Despite the differences in testing method, velocities used, and differences in product brands, the compiled data shows close groupings of filtration efficiency. Data on T-Shirt filtration in the present study is presented for both the lightweight and heavyweight t-Shirt.

**Figure 4:**
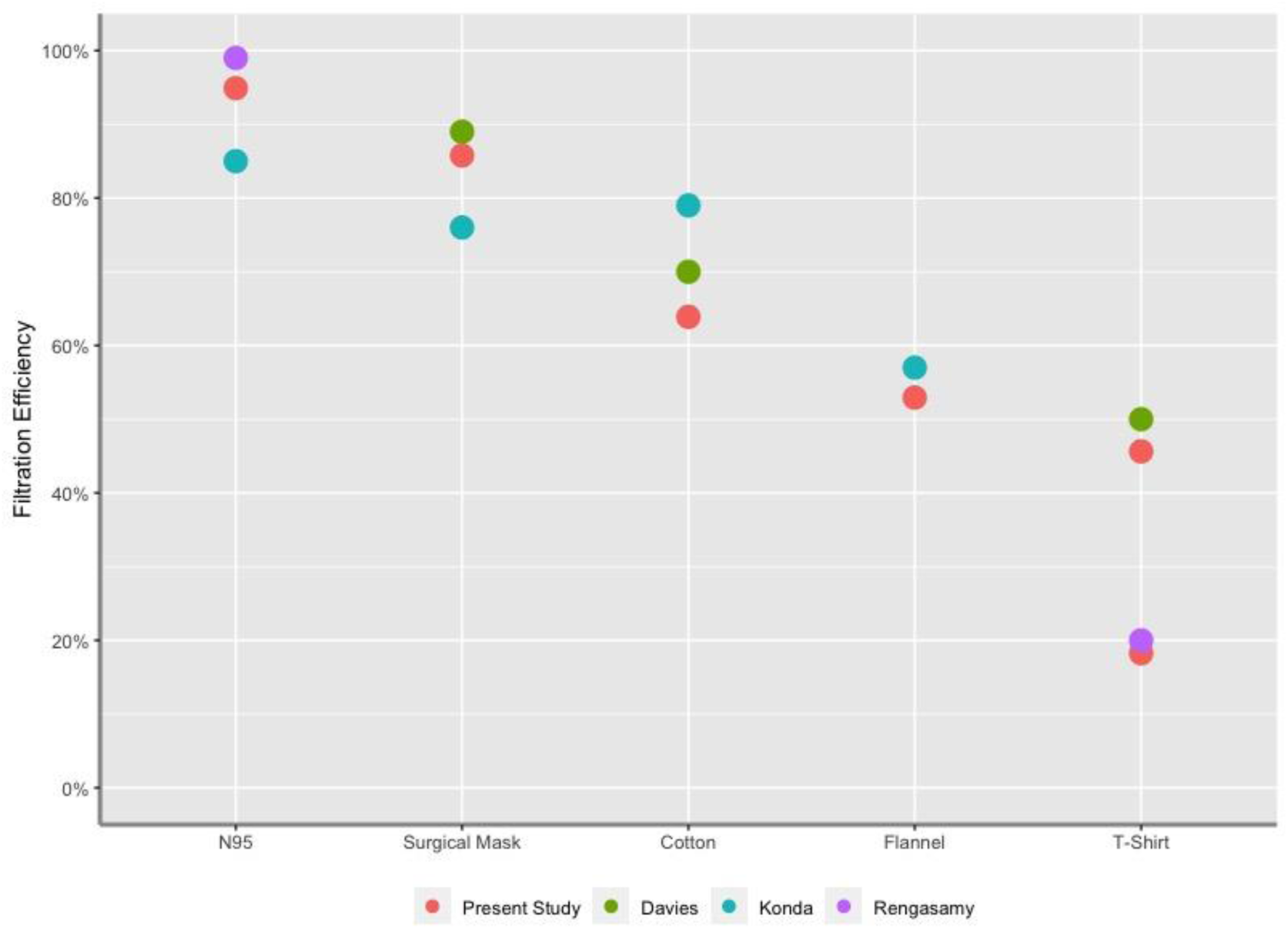
A comparison of existing data on fabric ultrafine filtration. Data chosen represents the highest velocity for each study. Data from this study was adjusted to proportionally represent a velocity of 0.2 m/s for this comparison. Data from Rengasamy et all is estimated from the included graphs, as statistical information about the data was not provided.

A comparison showed that no one study method consistently produced the highest results. Konda, who recorded the highest fabric FE also recorded the lowest FE for N95 and surgical masks. Surprisingly, Rengasamy et all’s data does not closely resemble Konda et all’s, although both studies compared NaCL filtration. This may be a factor of the Konda et all’s filtration studied at a greater velocity than Rengasamy et all uses, another indication for the importance of velocity on filtration. Our FE for fabric was frequently lower than others, a fact which may be accounted for with our single wash of the material before testing[11] and provide further evidence that washing fabric masks reduces their filtration efficiency.

### Safety Considerations

It is suggested homemade face masks should not be used in place of other protective measures such as self-isolation or social distancing. Rather, our results suggest homemade face masks may be a viable protective measure for those who cannot remain isolated and cannot obtain commercial face masks.

Repurposing material for homemade face masks comes with its own risks. Consideration should be given to respiratory hazards which may arise from the material used to construct a homemade facemask. For example, concern has been expressed that certain HEPA vacuum bags include fibers which, if inhaled, can cause lung injury. Lint and fibers from fabric, when inhaled in large quantities, and known to contribute to multiple lung problems including asthma, byssinosis, and bronchitis. For this reason, we would caution those needing to create homemade face masks to ensure all material is safe, nontoxic and lint-free. Fabrics which readily shed fibers may not be suited for face mask construction. The risks associated with such materials are an important area of further study, as large numbers of people are currently creating, wearing, washing, distributing, and selling homemade facemasks. Further research should further evaluate the ability of these materials and material combinations to filter specific viruses, pollutants, and other harmful airborne particles. Additional research on homemade facemask fit and fit testing is also critical at this time.

It is our hope that this study can assist home sewers and makers to create the best facemask possible when standardized commercial personal protective equipment is unavailable. Our study shows face masks can be created from common fabrics to provide wearers with significant protection from ultrafine particles. Until further research can establish the safety and viral filtration of fabric face masks, we suggest the use of approved respiratory protection whenever possible and the use of homemade face masks only when these products are unavailable.

It should be noted that the results of this study may also inform emergency mask creation in response to environmental emergencies where ultrafine particle levels are particularly dangerous, such as in the case of smoke or smog. Repeated face mask shortages during the California wildfires over the past few years have illustrated the recurring need for scientific data to guide the construction of homemade face masks when commercial supply chains are unable to meet demand.

## Data Availability

Data from this study is freely available under a CC-BY license on Cambridge University Apollo open data repository

https://doi.org/10.17863/CAM.51390

## AUTHOR’S CONTRIBUTIONS

### Eugenia O’Kelly

Conceived of the study, developed study methodology, obtained study materials and testing apparatus, collected study data, wrote manuscript

### Sophia Pirog

Obtained study materials, analyzed data and performed calculations, designed graphs, edited manuscript

### James Ward

Developed study methodology, reviewed data, edited manuscript, supervised study

### John Clarkson

Developed study methodology, reviewed data, edited manuscript, supervised study

## AWKNOWLEDGMENTS

The authors would like to thank Corinne E. O’Kelly for supporting this research. We would also like to thank the many sewers, doctors, and designers who spoke with us about their needs and concerns.

## DATA STATEMENT

Data from this study is freely available under a CC BY license on Cambridge University’s Apollo data repository.

**Link:** https://doi.org/10.17863/CAM.51390

**Citation:** O’Kelly, E., The Ability of Common Fabrics to Filter Ultrafine Particles [Data file]. Cambridge University: Cambridge, United Kingdom; 2020.https://doi.org/10.17863/CAM.51390

